# Resident Odor Reports and Differing Health Outcomes in Areas of Industrial Emission Odor, Louisville, Kentucky

**DOI:** 10.1101/2025.05.13.25327517

**Authors:** Angelina Rangel, Lauren B. Anderson, Rochelle H. Holm, Ted Smith

## Abstract

**Background:** Environmental odors can impact health and risk perception. Participatory, or citizen science, can provide important data through systematic, mobile application assisted odor reporting.

**Objectives:** This study leverages participatory public health science using the Smell MyCity app to investigate resident-reported odors and their association with health outcomes in Louisville, KY.

**Methods:** We analyzed 6,868 odor reports from 2018 to 2024, to identify census tracts where industrial and chemical odor reports cluster. Disease prevalence from the CDC’s PLACES data were compared between these tracts and the entire county.

**Results:** Results suggest associations between frequent odor reporting areas and increased health risks, highlighting the public health significance of environmental odors and the importance of community-driven data collection.

**Conclusions:** This community-driven reporting initiative may be a useful addition to health research in areas which also have high industrial emission odor.

## 1. Introduction

Environmental odors can affect health and influence perceptions of health risk.^1^ Community science is a powerful public health tool for odor reporting.^2^ While odors are typically regulated as a nuisance matter rather than as an indicator of air pollution exposure, ambient odors can degrade quality of life, causing symptoms ranging from headaches and gastrointestinal distress to respiratory issues.^3^ Due to the lack of standardized and timely data about ambient odors, the extent of these symptoms, other health effects, and the possibility that these odors are indicators of specific air toxics is difficult to ascertain.

## 2. Methods

The goals of the project were to: (1) analyze resident-reported odors and identify geographic patterns, (2) compare disease prevalence in areas with more frequent industrial and chemical resident odor reports to the rest of the county, and (3) showcase participatory public health science as a pioneering approach for prioritizing odor investigations by local authorities.

After a soft launch, the Smell MyCity app^4^ was introduced to the Louisville community in March of 2019 with support from community-based organizations, the University of Louisville, and local government agencies.^5^ The mobile phone applications prompt community members to rate the intensity of odors from 1 (just fine!) to 5 (almost as bad as it gets!), adding odor descriptions and other optional details and captures location information from the phone. Smell reports were immediately displayed on the public Smell MyCity map^4^, submitted to Louisville’s Air Pollution Control District and made available for public download. Our analysis focused on descriptions related to industrial and chemical odors, March 2018 through September 2024 (Supplementary Table S1). Coding was by two authors (AR and LBA). The ArcGIS Pro version 2.9.5 (Redlands, CA) Kernel Density tool was used to identify areas where noxious odor reports were highest across the county. We then compared disease prevalence rates from the 2022 PLACES data^6^ for asthma, cancer, chronic obstructive pulmonary disease (COPD), coronary heart disease (CHD), depression, diabetes, obesity, and stroke between the census tracts with higher odor reports and the countywide average.

The data analyzed were publicly available and therefore institutional review board approval was not required. All data were anonymous and do not include personal identifiers.

## 3. Results

The intervention was implemented in Louisville, Kentucky, a city with a long history of industrial odors and air pollution and a population of 793,881 people.^7^ Between 2018 and 2024, residents submitted 6,868 smell reports with odor descriptions to the Smell MyCity app; 1,119 reports described “industrial” and “chemical” odors and assigned smell values of 3 or greater. While odor reports were submitted from across Louisville, many reports originated in the northwestern area of the city. Kernel Density analysis highlighted two census tracts with high frequencies of industrial and chemical odor reports: Census Tract 14 (21111001400) with 163 reports and Census Tract 50 (21111005000) with 83 reports, in the Chickasaw (estimated population 6,299) and California (estimated population 6,523) neighborhoods respectively (Figure 1).^8^ Higher disease prevalence between the census tracts of interest and Louisville were observed for asthma, cancer, COPD, CHD, depression, diabetes, obesity, and stroke (Table 2). These neighborhoods are near the historically significant industrial corridor known as “Rubbertown” due to its prominence in the chemical production of synthetic rubber and related petrochemical activities. This area had previously been the focus of the West Louisville Air Toxics Study (WLATS) conducted between April 2000 and December 2005, a comprehensive air monitoring initiative in response to longstanding resident concerns about emissions from local industries. That research ultimately created the United States EPA Strategic Toxic Air Reduction (STAR) Program, which was locally enacted in June 2005, aimed to significantly reduce toxic air pollutants and address public health risks associated with industrial emissions in Louisville.^9^

**Figure 1.**
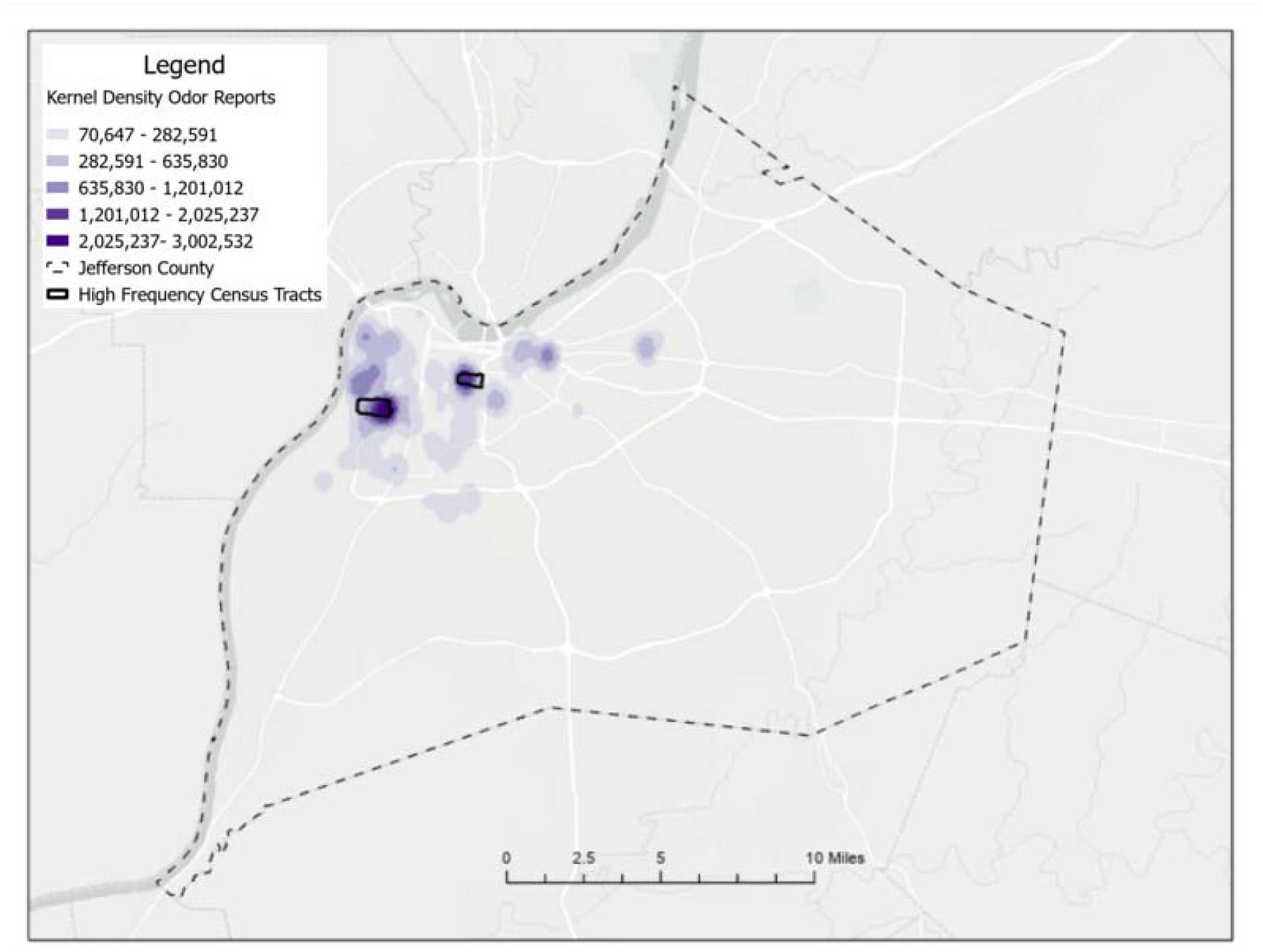
Map of Louisville, Kentucky: Census tracts with high odor report frequency are outlined in black. Kernel Density analysis highlighted two census tracts with high frequencies of industrial and chemical odor reports: Census Tract 14 (21111001400) with 163 complaints and Census Tract 50 (21111005000) with 83 complaints.

**Table 1.** Summary of mean 2022 PLACES disease prevalence rates for asthma, cancer, chronic obstructive pulmonary disease, coronary heart disease, depression, diabetes, obesity, and stroke for census tracts with a high density of odor reports compared to Louisville as a whole. 95% confidence intervals are provided for the countywide group.

| 2022 PLACES disease | High density odor report census tract 14 (21111001400) |  | High density odor report census tract 50 (21111005000) |  |
| --- | --- | --- | --- | --- |
|  | Census tract mean % | Countywide mean % (95% CI) | Census tract mean % | Countywide mean % (95% CI) |
| Asthma | 14.7% | 11.3% (11.1–11.5) | 13.5% | 11.3% (11.1–11.5) |
| Cancer | 4.8% | 7.9% (7.6–8.2) | 7.7% | 7.9% (7.6–8.2) |
| Chronic obstructive pulmonary disease | 11.1% | 9.2% (8.7–9.6) | 15.1% | 9.2% (8.7–9.6) |
| Coronary heart disease | 7.8% | 7.6% (7.3–7.8) | 11.9% | 7.6% (7.3–7.8) |
| Depression | 24.3% | 25.6% (25.3–25.9) | 26.9% | 25.6% (25.3–25.9) |
| Diabetes | 21.7% | 14.3% (13.7–14.9) | 21.8% | 14.3% (13.7–14.9) |
| Obesity | 51.7% | 37.9% (36.9–38.8) | 44.0% | 37.9% (36.9–38.8) |
| Stroke | 6.4% | 4.1% (3.8–4.3) | 7.7% | 4.1% (3.8–4.3) |

The members of the West Jefferson County Community Task Force (WJCTF) and Rubbertown Emergency Action (REACT) have advocated for cleaner air, monitored air quality, and assessed health risks associated with air pollutants for residents living in west Louisville. The WJCCTF and REACT promoted the Smell MyCity initiative with the aim to enhance transparency in reporting, bolster citizen trust, and effectively collect data on problematic areas to bring to the attention of APCD.

## 4. Discussion

The motivation behind this intervention was to re-valuate the utility of odors, especially those near industrial sources, as possible indicators of environmental health risks rather than limiting their value as a public nuisance as characterized by air quality regulation. By exploring associations between places with frequent industrial and chemical odor reports and observed disease burden, a new seriousness may be given to the frequent presence of these odors. Furthermore, by empowering residents to report their observations in such a participatory science framework, communities can more directly affect public perception and policy.^10^ Thus, our project aimed to investigate the association between industrial and chemical odor reports and the prevalence of specific health conditions, expanding data sources which may be useful to identify and address potential health disparities. We aimed to determine whether there is an association between community-reported odors and a range of health conditions in an area with industrial emissions for prioritizing investigations by local authorities.

Continuing the use and promotion of applications like Smell MyCity can empower residents to document and report odors, thereby enabling detailed investigations into clusters of odor sources and related health impacts. This not only improves public health but also strengthens environmental justice initiatives. However, sustainability ultimately depends on ongoing application maintenance by third-party providers, such as Smell MyCity’s Carnegie Mellon team, and ensuring the app remains free for the public. Unlike other research that aligns odor reports with major spatial landmarks for prioritization,^11^ our intervention uniquely integrates these reports with health outcomes to enhance prioritization strategies. The sustainability of this activity is facilitated by strong collaborations among community organizations, academic institutions, and local government agencies committed to analyzing the data and engaging. These partnerships foster resource sharing and collaborative problem-solving, driving active community involvement and data-driven public health advocacy.

## 5. Conclusion

Research and analysis of participatory community data, made possible through programs like Smell MyCity, play a crucial role in preserving trust and engagement in public health activities at both local and broader scales. Ultimately, our approach offers a potential model for how grassroots odor reporting efforts can be recognized and respected in environmental health and policy, benefiting both local communities and broader public health initiatives.

## Data Availability

Smell MyCity source data are publicly available, links are provided in the references.

## ACKNOWLEDGMENTS

We would like to extend our sincere thanks to the Carnegie Mellon CREATE Lab for developing the Smell MyCity smartphone application and for making its data freely available to the public. We also thank Anna Dusterhoff for data support. This work was supported by the Owsley Brown II Family Foundation and the NIH (P42 ES023716). The funders had no role in the study design, data collection and analysis, decision to publish, or manuscript preparation.

## COMPETING INTERESTS

The authors declare that they have no known competing financial interests or personal relationships that could have appeared to influence the work reported in this paper.

## CONTRIBUTORS

Conceptualization: TS; Methodology: LBA, TS; Formal analysis: AR, LBA; Writing-original draft preparation: LBA, RHH; Writing-review and editing: AR, LBA, RHH, TS; Supervision: TS; Project administration: LBA. All the authors have read and agreed to the published version of this manuscript.

## DATA SHARING

Smell MyCity source data are publicly available, links are provided in the references.

## Supplementary Material

**Supplementary Table S1.**
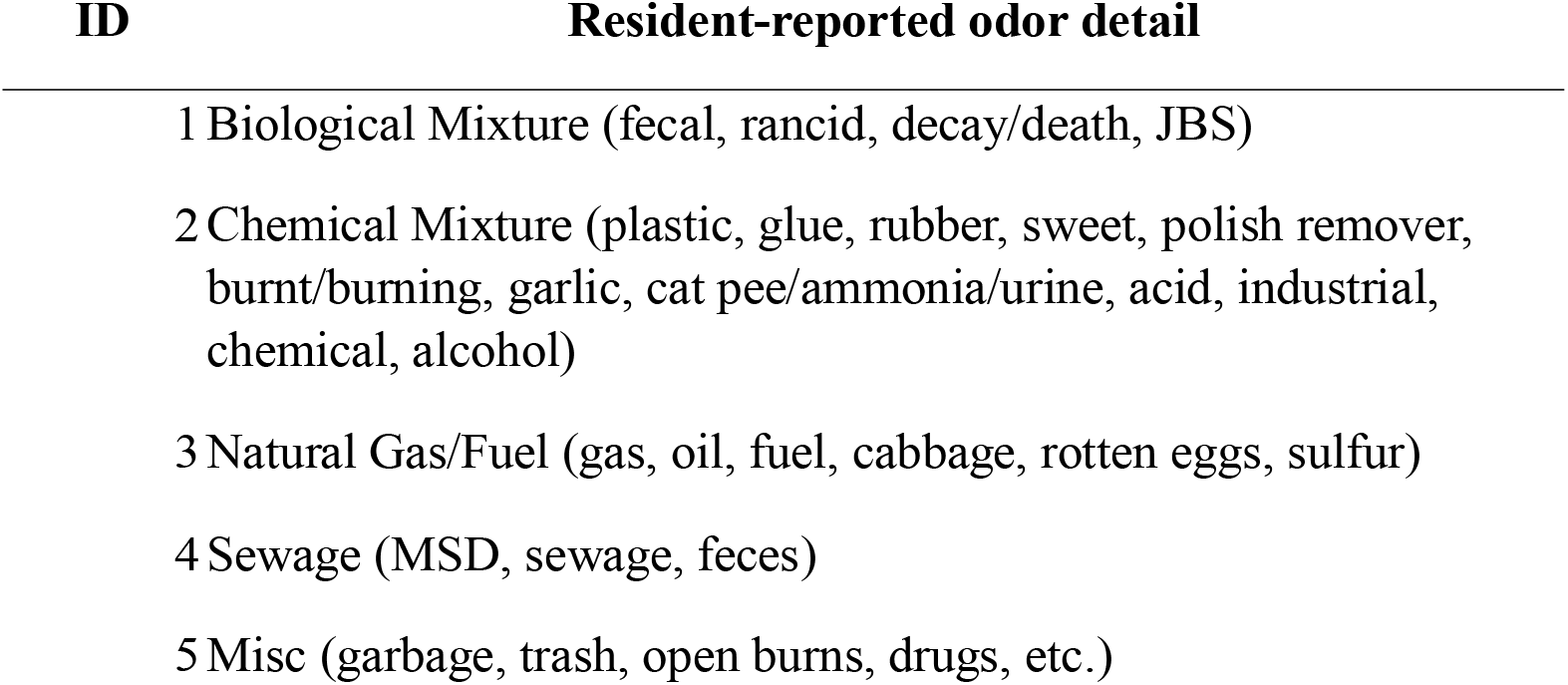
Code analysis for reported odors from the Smell MyCity app. This study was limited to odor descriptions related to industrial and chemical odors (ID 2).

| **ID** | **Resident-reported odor detail** |
| --- | --- |
| 1 | Biological Mixture (fecal, rancid, decay/death, JBS) |
| 2 | Chemical Mixture (plastic, glue, rubber, sweet, polish remover, burnt/burning, garlic, cat pee/ammonia/urine, acid, industrial, chemical, alcohol) |
| 3 | Natural Gas/Fuel (gas, oil, fuel, cabbage, rotten eggs, sulfur) |
| 4 | Sewage (MSD, sewage, feces) |
| 5 | Misc (garbage, trash, open burns, drugs, etc.) |

## Notes

### Competing Interest Statement

The authors have declared no competing interest.

### Summary of Updates

Updated to correct minor grammatical mistakes.

